# Predicting Hospital Readmission among Patients with Sepsis using Clinical and Wearable Data

**DOI:** 10.1101/2023.04.10.23288368

**Authors:** Fatemeh Amrollahi, Supreeth Prajwal Shashikumar, Haben Yhdego, Arshia Nayebnazar, Nathan Yung, Gabriel Wardi, Shamim Nemati

**Affiliations:** Division of Biomedical Informatics, University of California San Diego, La Jolla, CA 92093; Department of Emergency Medicine, UC San Diego Health, La Jolla, CA 92093; Division of Pulmonary, Critical Care and Sleep Medicine, UC San Diego Health, La Jolla, CA 92093

**Keywords:** Sepsis, Activity level, Hospital readmission, Wearable data

## Abstract

Sepsis is a life-threatening condition that occurs due to a dysregulated host response to infection. Recent data demonstrate that patients with sepsis have a significantly higher readmission risk than other common conditions, such as heart failure, pneumonia and myocardial infarction and associated economic burden. Prior studies have demonstrated an association between a patient’s physical activity levels and readmission risk. In this study, we show that distribution of activity level prior and post-discharge among patients with sepsis are predictive of unplanned rehospitalization in 90 days (P-value*<*1e-3). Our preliminary results indicate that integrating Fitbit data with clinical measurements may improve model performance on predicting 90 days readmission.

## I. INTRODUCTION

Sepsis is defined as a dysregulated host response to infection and remains a major public health burden [1]. Worldwide, approximately 50 million people develop sepsis each year and it is estimated that sepsis is responsible for 1 out of every 5 global deaths [2]. In the United States, it is estimated that over 1.7 million adults in America develop sepsis every year, resulting in over 350,000 deaths [3].

While there have been significant efforts to decrease mortality of sepsis, recent data demonstrate that sepsis survivors are at high risk of various complications, including prolonged immunosuppression, physical deconditioning and neurocognitive impairments [4]. This may explain why patients with sepsis have significantly higher rates of unplanned readmissions than patients with heart failure, myocardial infarction, pneumonia and chronic obstructive lung disease (COPD) [5], [6], [7]. Additionally, recent data suggests sepsis readmissions are responsible for over $3.5 billion dollars a year in healthcare expenditures in the United States alone [6]. Importantly, there have been only a handful of interventions that have shown promise to decrease unplanned sepsis readmissions [8]. The importance of care after discharge cannot be understated. A 10-year Taiwanese study of over 15,000 patients found a 5.6% mortality risk reduction in those who received rehabilitation within 3 months of discharge [9]. A German study found decreased mortality among patients who suffered from a severe infection or more and were sent to post-discharge rehabilitation [10].

Wearable devices such as Fitbit allow for tracking vital signs and physical activity levels have proliferated continuously in the past decades, but the involvement of wearable data within clinical applications remains scarce [11].

Nemati et al. developed a model to detect atrial fibrillation using photoplethysmogram (PPG) recorded from simband smartwatches with an area under the receiver operating curve (AUROC) of 0.99 [12]. W. J.Kane et al. used step count recorded by wearable devices and showed that 10% increase in step count 30 days after colorectal surgery decreased the risk of 30 days readmission by 40% [13]. Such devices may help identify patients at high risk for physiologic deterioration and potential readmission to the hospital. In this study, we hypothesized that wearable data are predictive factors for assessing the risk of rehospitalization among patients with sepsis.

## II. METHODS

### A. Population Study

We conducted a multicenter retrospective study using data available from the AllofUS (C2021Q3R6) data repository [14]. AllofUs contains electronic health records (EHR) of over 372,380 participants from May 2018 to June 2022. Institutional Reviewing Board (IRB) approval was obtained prior to enrollment of patients in the AllofUs Research Program, the data has been deidentified, and has been made available in a secure enclave for research purposes.

We included all patients 18 years or older admitted to the hospital for at least two days that had sepsis, which was determined according to the Sepsis-3 criteria, following our previous work on this dataset [15]. We excluded patients who expired or were transitioned to hospice during their admission, or died after hospital discharge. In addition, we excluded patients encountered without any measurement of vital signs. For patients with multiple hospitalizations for sepsis, we included the first, or “index” sepsis admission and corresponding 90-day readmission, if present.

### B. Preprocessing and Features

Patient clinical and physiological data during their hospital stay as well as Fitbit data pre- and post-discharge following 90 days after their discharge or until their readmission time were used in our analysis. A total of 40 physiological and clinical variables well established by prior sepsis studies were collected for each patient during their hospitalization [15], [16], [17], [18]. We included pre-discharge and post-discharge wearable data including heart rate, basal metabolic rate calories and activity levels averaged per day. For the analysis we utilize 5, 50, and 95 percentile of each measurement. Missing values of clinical and continuous variables were imputed using mean estimated values and normalized. We used one hot encoding and KNN imputation for the remainder of binary and categorical features.

### C. Model Development and Validation

Each model was evaluated using subject-wise leave-one-out cross validation. Area under the receiver operator characteristic curve (AUCroc), specificity and positive predictive value at threshold corresponding to 80% sensitivity were used as evaluation metrics.

To evaluate the role of integrating pre-discharge and post-discharge wearable data we compared six distinct models that utilize different combinations of input features. We used random forest classifiers with 19 trees and 4 minimum samples to split internal nodes. We used model interpretability methods (namely Shapley values) to rank the importance of the different features. Additionally, we assessed the change in feature importance as a function of time since hospital discharge.

## III. RESULTS & DISCUSSION

We identified 9,910 index admissions with sepsis, among which 2815 were readmitted in 90 days (29%). Table I summarizes characteristics of patients who were readmitted and not readmitted within 90 days of hospital discharge.

**TABLE I.**
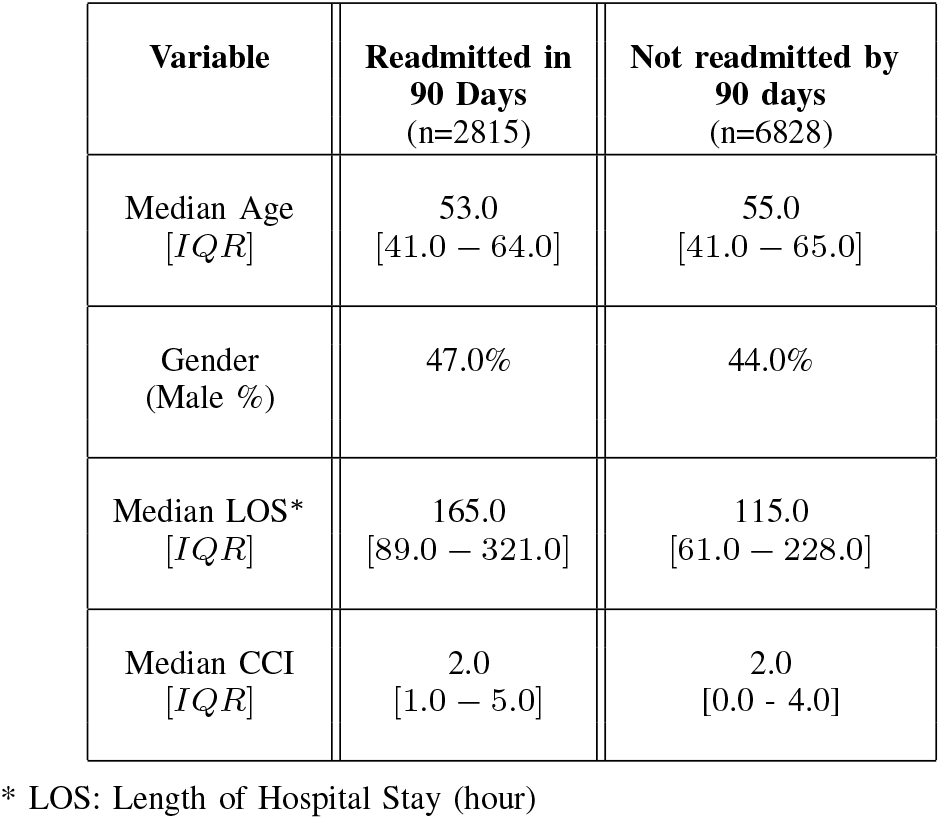
Patients characteristics

| Variable | Readmitted in 90 Days<br>(n=2815) | Not readmitted by 90 days<br>(n=6828) |
| --- | --- | --- |
| Median Age<br>[IQR] | 53.0<br>[41.0 – 64.0] | 55.0<br>[41.0 – 65.0] |
| Gender<br>(Male %) | 47.0% | 44.0% |
| Median LOS*<br>[IQR] | 165.0<br>[89.0 – 321.0] | 115.0<br>[61.0 – 228.0] |
| Median CCI<br>[IQR] | 2.0<br>[1.0 – 5.0] | 2.0<br>[0.0 – 4.0] |

\* LOS: Length of Hospital Stay (hour)

We identified 103 patients in the current release of the AllofUS dataset (2021Q3R6) with Fitbit recordings. Of these, thirty-nine of patients with sepsis had Fitbit data available during their hospitalization (pre-discharge records) or after hospital discharge (post-discharge records), 7 of whom were readmitted in 90 days. Figure 1 shows the association of sedentary hours per day and readmission risk. We found that patients who were more sedentary were more likely to be readmitted by 90-days post-discharge. Table II provides each model’s input and their performances.

**TABLE II.** Comparison of Model Performances

| Model<br>(input features) | AUCroc | Specificity* | PPV* |
| --- | --- | --- | --- |
| Model I:<br>(Pre-discharge clinical<br>variables) | 0.74 | 0.47 | 0.26 |
| Model II:<br>(Pre-discharge wearable<br>data) | 0.6 | 0.34 | 0.25 |
| Model III:<br>(Post-discharge wearable<br>data) | 0.68 | 0.56 | 0.3 |
| Model IV:<br>(Pre-discharge Clinical<br>Predischarge wearable) | 0.78 | 0.43 | 0.25 |
| Model V:<br>(Pre-discharge<br>Post-discharge wearable) | 0.80 | 0.56 | 0.3 |
| Model VI:<br>(Clinical Variables, Pre-discharge<br>& Post-discharge wearable) | 0.85 | 0.87 | 0.6 |

\* Measured at 80% sensitivity

**Fig. 1.**
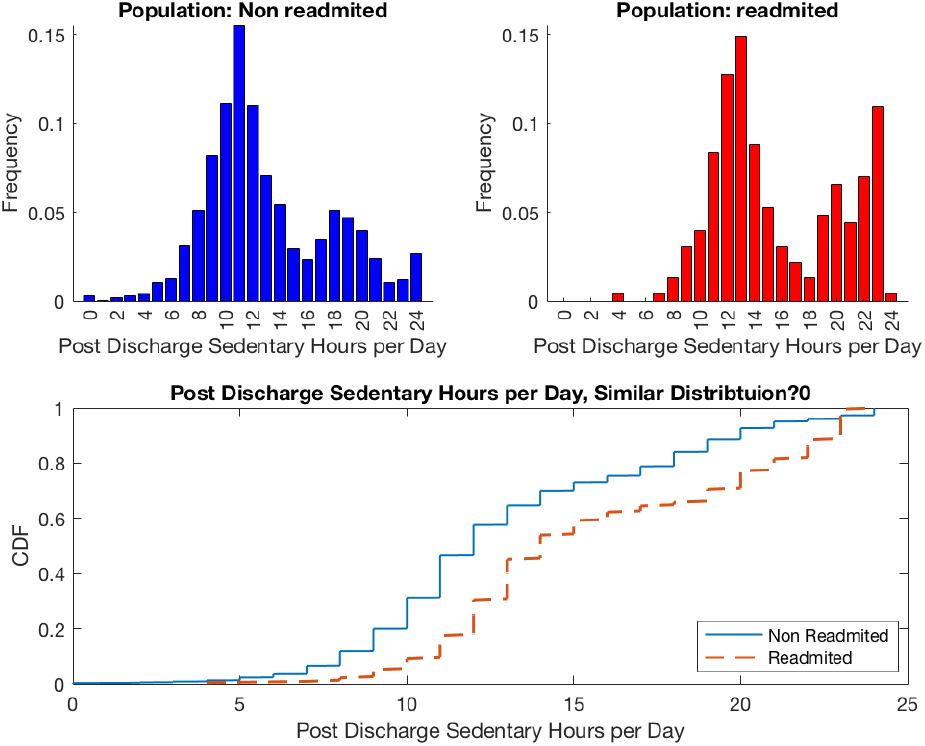
An illustrative example of assessing feature distribution among patients with sepsis re-admitted vs non readmitted. Upper panels show the histogram of the sedentary hours of every day, recorded every minute, for readmitted vs non readmitted patients with sepsis. The lower panel shows the CDF for both population cohorts and the kolmogorov–Smirnov test (KS test) reveals that the distribution of activity hours for re-admitted vs non readmitted patients are dissimilar.

We found that Model 6, which included clinical data as well as pre- and post-discharge Fitbit data had the best ability to predict a readmission, including a PPV of 60% at 80% sensitivity. In comparison, Model 1 which only included pre-discharge clinical features had a PPV of 26% at 80% sensitivity.

Figure 2 illustrates top 5 predictor factors for the risk of readmission at day 1 and day 10 post-discharge. We found that at day 1 pre-discharge features such as Sedentary rate, *O*_2_*Sat*, blood Glucose level, CRP (a biomarker of inflammation) and heart rate were significant predictors of readmission. By day 10, factors such as post-discharge Sedentary rate showed significant predictive value.

**Fig. 2.**
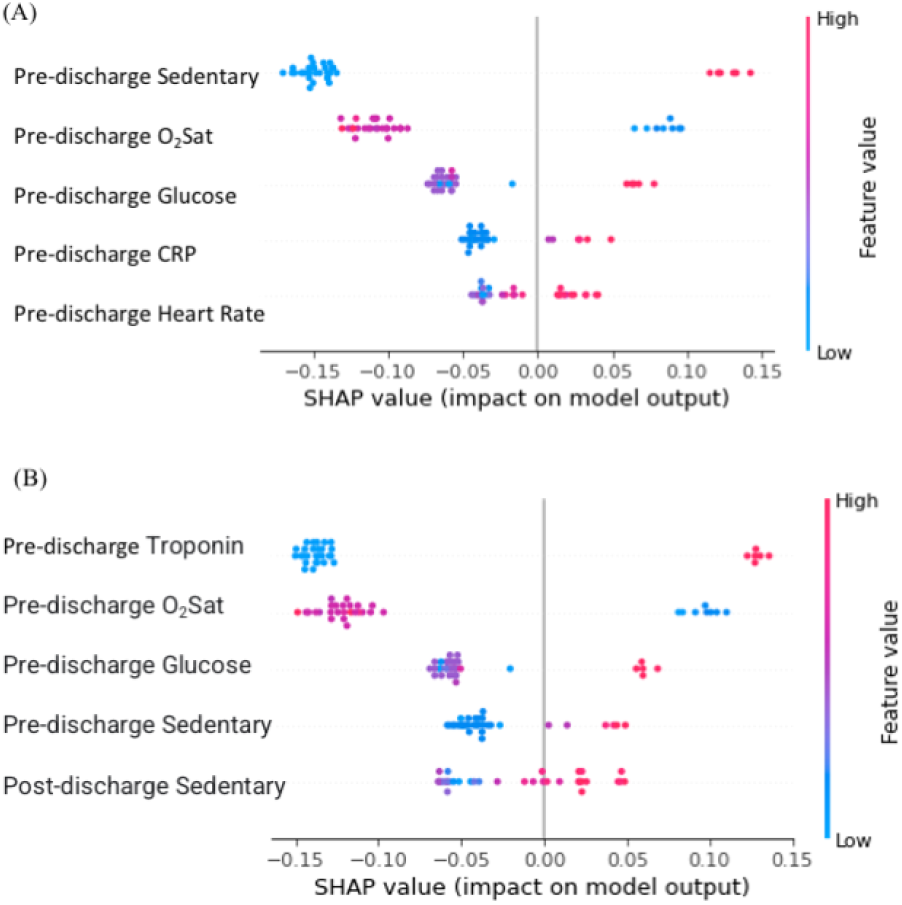
Shapley values of the top 5 predictive features in readmission risk. (A): Post discharge day 1 (B): Post discharge day 10. CRP = C-reactive protein, Sedentary= Sedentary minutes per day (Activity level).

There are few validated models that accurately predict readmission following an index hospitalization for sepsis. Commonly used tools, such as the LACE+ index and HOSPITAL score were not developed specifically for sepsis patients and perform poorly at predicting readmissions in this population [19]. A previous EHR-based model developed by our team [19], using only pre-discharge clinical features, demonstrated improved predictive power over LACE+, but the model AUCroc was around 0.71. An accurate model for assessing the risk of readmission may help healthcare providers allocate resources to higher risk patients and prevent unplanned readmissions.

While in this work we focused on readmission prediction, equally important is the problem of assessing discharge readiness, or more broadly figuring out if de-escalation of care-level is warranted. A comparison of Model II and Model IV indicate that wearable data are likely to contribute to de-escalation of care and discharge planning. Moreover, effective implementation of preventive strategies for reduction of unplanned readmission often requires involving a care-coordinator three or more days in advance of patient discharge. As noted by Kash1 and colleagues [20], some of the most effective interventions to reduce unplanned readmissions include better coordination among clinical teams and/or community providers, post-discharge home visits, telephone follow-up calls, patient/family education, and discharge planning. As such, continuous prediction of discharge readiness and prediction of unplanned readmission during hospitalization is likely to help with timely care-coordination in order to avoid unplanned readmission.

Fitbit or other wearable data can also provide monitoring for sedentary behaviors in the after-discharge period which is known to also be associated with increased mortality. On a national level, the Centers for Medicare and Medical Services (CMS) uses the rehospitalization rate to adjust the hospitals’ reimbursement in favor of hospitals with a lower readmission rate [21].

Additionally, sepsis readmissions carry a major economic burden and effective strategies to curtail this are lacking. Although our data are small, our preliminary results suggest there may be benefit of incorporating Fitbit data (or other wearable devices) into risk prediction models to decrease the economic burden of sepsis readmissions. This work developed different models for predicting the risk of readmission for patients with sepsis based on their clinical and wearable data before and after discharge. Our major finding is that Model 6 (which uses clinical features, including pre-discharge and post-discharge wearable data) significantly outperformed the other models in all performance metrics. Importantly, we found that the model trained with post-discharge wearable data features had higher specificity and PPV than models that trained on other input features. We acknowledge several limitations to our work. First, data were retrospective and we are unable to demonstrate causation. Next, we only had a small number of patients with Fitbit data which limits our findings.

In summary, we found that models using wearable sensor data may have superior performance in predicting 90-day readmissions following an index sepsis hospitalization than those that do not utilize this data. Future, prospective studies should be conducted to evaluate the performance of these models.

## Data Availability

We conducted a multicenter retrospective study using data available from the AllofUS (C2021Q3R6) data repository

## Acknowledgment

S.N. is funded by the National Institutes of Health (R01LM013998, R01HL157985, R35GM143121). He is co-founder of a UCSD start-up, Healcisio Inc., which is focused on commercialization of advanced analytical decision support tools. G. W. is funded through an early career award from the National Institute of General Medical Sciences (K23GM146092).

## Notes

### Competing Interest Statement

The authors have declared no competing interest.

### Funding Statement

S.N. is funded by the National Institutes of Health (R01LM013998, R01HL157985, R35GM143121). S.N. and S.P.S are cofounders of a UCSD start-up, Healcisio Inc., which is focused on commercialization of advanced analytical decision support tools. G. W. is funded through an early career award from the National Institute of General Medical Sciences (K23GM146092).

### Author Declarations

Institutional Reviewing Board (IRB) approval was obtained prior to enrollment of patients in the AllofUs Research Program, the data has been deidentified, and has been made available in a secure enclave for research purposes.

